# Normal appearing white matter and disability in multiple sclerosis

**DOI:** 10.1101/2025.11.07.25339773

**Authors:** Valentin N. Stepanov, Santiago Coelho, Benjamin Ades-Aron, Jenny Chen, Timothy M. Shepherd, Dmitry S. Novikov, Ilya Kister, Els Fieremans

## Abstract

**Background and Objectives:** Multiple sclerosis (MS) is characterized by lesions and atrophy on conventional MRI, yet these often fail to explain disability. Diffusion MRI (dMRI) detects microstructural injury with diffusion tensor (DTI) and kurtosis imaging (DKI) offering sensitivity, and Standard Model Imaging (SMI) providing biologically interpretable parameters. We evaluated whether these clinically feasible dMRI metrics are associated with disability beyond volumetric and lesion measures, and whether effects arise from normal-appearing white matter (NAWM).

**Methods:** This cross-sectional study included MS patients who underwent 3T MRI including T1- and T2-weighted and a ∼7-minute multi-shell dMRI protocol. Brain volumes (gray matter, white matter, thalamus) and lesion load were derived using FreeSurfer and Icobrain. Diffusion metrics included radial diffusivity (RD) from DTI, radial kurtosis (RK) from DKI, intra-axonal water fraction (*f*) and fiber dispersion (*p_2_*) from SMI. Clinical outcomes were the Expanded Disability Status Scale (EDSS), Multiple Sclerosis Functional Composite (MSFC), 9-Hole Peg Test (9HPT), Symbol Digit Modalities Test (SDMT), and disease duration. Voxelwise and tract-based regions of interest analyses were adjusted for sex and age at onset and repeated after excluding lesions.

**Results:** Ninety-two patients (68 women; mean age 48, range 24–73; median disease duration 14 years; EDSS 3.0, range 0-8.5) were included. dMRI revealed widespread associations with all clinical measures that persisted after lesion exclusion, implicating NAWM. Functional outcomes were tract-specific: 9HPT correlated with corticospinal tract and optic radiations (RD ρ=0.45; RK ρ=–0.44; *f* ρ=–0.42); MSFC with brainstem and optic radiations (RD ρ=–0.52; RK ρ=0.40; *f* ρ=0.39). SDMT showed widespread correlation with diffusion and atrophy (white matter ρ=0.49; thalamus ρ=0.47). EDSS showed weaker diffusion highlighting commissural disorganization (forceps major/minor ρ ≈– 0.30 to –0.32) and was most strongly associated to infratentorial lesion load (ρ=0.42). Disease duration was dominated by gray-matter atrophy (ρ=–0.54) with commissural *p_2_* reductions (≈–0.45).

**Discussion:** dMRI detects NAWM injury underlying functional impairment beyond atrophy and lesions. SMI adds specificity (*f* for axonal loss/demyelination; *p_2_* for inflammation). Structural measures capture the effect of cumulative burden in terms of disease duration and EDSS. Together, diffusion, volumetric, and lesion metrics offer complementary insights, supporting multimodal imaging for MS monitoring and stratification.

## INTRODUCTION

Multiple Sclerosis (MS) is a disease of the central nervous system characterized by inflammation, demyelination, and axonal injury ^1^. On conventional T1- and T2-weighted MRI, MS is primarily identified by white matter (WM) and gray matter (GM) lesions ^2–4^, which combined with atrophy, improve diagnosis and monitoring ^2^. Yet atrophy reflects late-stage neurodegeneration and often misses early pathology ^1,5^. Likewise, LL does not fully explain disability, as patients with similar lesion burden can differ markedly in severity ^6^. Together these caveats underlie the “clinico-radiological paradox,” highlighting the need for imaging methods that detect subtle injury and better predict outcomes.

Diffusion MRI (dMRI) probes the random microscopic motion of water molecules within tissue, capturing their overall diffusion displacement term, known as the propagator^7^. It is highly sensitive to microstructural damage in the normal-appearing WM (NAWM) invisible to structural MRI^8^. dMRI quantification is conventionally done using Diffusion Tensor Imaging (DTI), which characterizes Gaussian diffusion, enabling anisotropy measurement, tract visualization, and detection of NAWM abnormalities ^9–11^. Diffusion Kurtosis Imaging (DKI) extends this by quantifying additional complexity from non-Gaussian diffusion^12,13^.

However, both DTI and DKI lack direct specificity to pathology. Biophysical modeling addresses this limitation by estimating biologically interpretable parameters from dMRI. In WM, the Standard Model (SM) ^7,14^ describes intra- and extra-axonal compartments together with an orientation distribution function (ODF). Standard Model Imaging (SMI) extracts these parameters from clinically feasible two-shell data ^15–17^, including the intra-axonal water fraction, *f*, i.e. the relative contribution of the intra- and extra-axonal water (excluding myelin), and *p*_2_, the 2^nd^ order rotational invariant representing the fiber orientation distribution function. By design, SM parameters provide higher specificity to cellular pathology, where both demyelination and axonal loss will cause a decrease in *f*, and gliosis e.g. by activated microglia and astrocytosis will increase dispersion and cause a decrease in *p*_2._ SM parameters have been validated against histology in animal models ^18,19^ and realistic simulations ^20,21^. Recent studies in MS demonstrate that SMI metrics capture disability across disease stages (mild vs severe disability)^8^, underscoring their clinical relevance for stratification and progression tracking.

Systematic comparisons between DTI/DKI, LL, and atrophy in relation to disability remain limited. Here, we analyze damage in global WM and NAWM using DTI, DKI, and SMI alongside volumetric and lesion-based measures. Our goal is to determine whether quantitative dMRI metrics (DTI/DKI/SMI) provide added value over brain volume and lesion load in tracking disability. We hypothesize that (i) dMRI will enhance sensitivity to early and diffuse alterations complementing atrophy; (ii) SMI provides specificity to cellular pathology; and (iii) NAWM pathology drives much of the clinical variance. We apply voxel-wise and a region-of-interest (ROI) analyses spanning major WM pathways to test whether spatially specific microstructural patterns explain clinical outcomes. We also assess the impact of lesion exclusion to determine the extent to which NAWM damage contributes to disease progression. Overall, our study aims to determine whether advanced, yet clinically feasible, dMRI can serve as a robust and sensitive marker of disease progression, informing therapeutic decisions and clarifying MS pathology.

## METHODS

### Subjects

This study was approved by the local Institutional Review Board, and all participants provided written informed consent prior to inclusion. A total of 128 patients were initially recruited from the NYU Langone Multiple Sclerosis Comprehensive Care Center (New York) between October 2021 and December 2024. Of these, 28 withdrew before MRI scanning due to clinical deterioration, transportation difficulties, or personal circumstances, leaving 100 who underwent MRI and clinical evaluations. Of the 100 patients, 8 were excluded due to incomplete MRI acquisitions, resulting in a final cohort of 92 patients (68 females, 24 males; mean age 48 years, range 24-73). Their demographic and clinical characteristics are summarized in Table 1. All patients were diagnosed with relapsing or progressive MS by a neuroimmunologist in accordance with 2017 revision of McDonald criteria ^3^ and had been relapse-free for at least three months prior to enrollment. Most of the participants were on disease-modifying therapy at the time of the study. All clinical evaluations were performed by neurologists specializing in MS.

**Table 1.** Demographic, clinical and MRI characteristics of the study cohort.

|  | **All patients**<br>**(n=92)** |
| --- | --- |
| **Sex** |  |
| *Female, n (%)* | 68 (74) |
| *Male, n (%)* | 24 (26) |
| **Mean age, years, mean (range)** | 48 (24-73) |
| **Race/ethnicity** |  |
| *White, n (%)* | 37 (40) |
| *Black, n (%)* | 37 (40) |
| *Hispanic, n (%)* | 14 (15) |
| *Other, n (%)* | 1 (1) |
| *Unknown, n (%)* | 3 (3) |
| **Median disease duration, years (range)** | 14 (0-45) |
| **EDSS, median (range)** | 3 (0-8.5) |
| **Flair brain lesion load, cm<sup>3</sup>, median (range)** | 4.7 (0.1-32.2) |
| **Brain total gray matter fraction at baseline %, median (range)** | 0.40 (0.32-0.54) |
| **Brain total white matter fraction at baseline %, mean (range)** | 0.29 (0.13-0.38) |

### Clinical Assessments

Clinical metrics included the Expanded Disability Status Scale (EDSS), Symbol Digit Modalities Test (SDMT), and Multiple Sclerosis Functional Composite (MSFC). EDSS is used to quantify disability in MS, with a focus on ambulation and neurological function ^22^. SDMT assesses cognitive processing speed ^23^, while MSFC is a composite measure evaluating cognitive and motor functions ^24^.

### Image Acquisition

All participants underwent brain MRI on a 3T scanner (Siemens Prisma, Siemens Healthineers, Erlangen, Germany) using a 20-channel phased array RF coil for reception. The MRI protocol included both structural and diffusion-weighted sequences.

Anatomical MRI was performed using a magnetization-prepared rapid gradient-echo (MPRAGE) (repetition time (TR) = 2300 ms, inversion time (TI) = 900 ms, echo time (TE) = 2.98 ms, flip angle (FA) = 9°, matrix = 256×256, voxel size = 1 mm isotropic, the total acquisition time (TA) = 5min 17s). In addition, a 3D fluid-attenuated inversion recovery (FLAIR) sequence was acquired to evaluate white matter lesions (TR = 5000 ms, TI = 1800 ms, TE = 393 ms, flip angle = 120°, matrix = 256×256, and voxel size = 1.0 mm isotropic, TA = 5min 23s).

Diffusion MRI (dMRI) was acquired using a spin-echo echo-planar imaging (EPI) sequence with the following parameters: TR = 4300 ms, TE = 92 ms, FA = 90, 68 slices, matrix size = 110×110, voxel size = 2 mm isotropic. Diffusion encoding was performed along directions uniformly distributed on a sphere at b = 0 (7 directions), b = 1000 (25 directions), and b = 2000 (60 directions) s/mm². Additional parameters included a multiband factor (MB) = 2, parallel imaging acceleration (GRAPPA) = 2, and partial Fourier acquisition = 6/8, TA ∼ 7 minutes.

### Image processing

#### Structural volume

Cortical, subcortical, and WM lesion segmentations were performed on T1-weighted MP-RAGE and FLAIR images using FreeSurfer v7.4.1 ^25^ with Samseg ^26^, and also using the commercial tool Icobrain ms v5.14.0 (Icometrix, Leuven, Belgium). From FreeSurfer, the following volumes were extracted: total WM, total gray matter (GM), subcortical GM (sGM), thalamus, and total lesion load (LL). Each FreeSurfer-derived volume, including LL was normalized to the subject’s total intracranial volume (TIV) ^27^.

Using Icobrain ms, volumes for total WM, total GM, total lesion load (LL), juxtacortical lesions (LL jxtc), infratentorial lesions (LL infratent), periventricular lesions (LL perivent), and deep WM lesions (LL deep wm) were obtained. Tissue volumes in Icobrain are normalized for head size by multiplying with a scaling factor derived from the affine transformation between MNI space and the individual T1 image, which reflects global volume change. All segmentations were visually inspected by a neuroradiologist with 7 years of experience (VS).

#### Diffusion metrics

dMRI data was pre-processed using the DESIGNER v2 pipeline (Python implementation) (https://github.com/NYU-DiffusionMRI/DESIGNER-v2) ^15,28^, including denoising ^29,30^, Gibbs artifact correction, EPI-induced distortion correction, motion and eddy current correction ^31^, and Rician noise floor correction. Parametric maps were estimated using the *tmi* tool, part of the DESIGNER v2 toolbox (https://nyu-diffusionmri.github.io/DESIGNER-v2/docs/TMI).

The diffusion tensor and kurtosis tensor were estimated using weighted linear least squares (WLLS) estimation ^32^. Note that kurtosis metrics were derived from the fourth-order cumulant tensor W (W maps) by taking respective traces ^33^. Parametric maps included mean diffusivity (MD), axial diffusivity (AD), radial diffusivity (RD), fractional anisotropy (FA), mean kurtosis (MK), axial kurtosis (AK), and radial kurtosis (RK). We also extracted SMI maps ^15^ of the intra-axonal volume fraction (*f*) and orientation dispersion index (*p_2_*), as these are the two most reliable SM metrics that can be estimated from two-shell dMRI protocols ^17^.

#### Voxel-Wise Analysis

Voxel-wise statistical analysis was performed using a non-parametric permutation-based approach ^34^. dMRI and SMI parametric maps were aligned to a multicontrast population-specific template, generated by combining all subjects’ FA and MD maps ^35^. This template allowed for the registration of all subjects into a common space. White matter was masked by FA > 0.2. Permutation testing was conducted with 5000 iterations, using Threshold-Free Cluster Enhancement (TFCE) to identify significant clusters ^35^.

Corrections for multiple comparisons were applied, with a family-wise error (FWE) rate of p ≤ 0.05 considered significant. Voxel-wise correlations between diffusion metrics and clinical measures - 9HPT, MSFC, EDSS, SDMT, and disease duration (DD) were performed. Correlations were calculated while controlling for sex and age at disease onset.

#### ROI Analysis

ROI analysis was conducted using TractSeg ^36^ to perform tractography-based segmentation of major white matter tracts directly in subject space, eliminating the need for inter-subject registration. To increase segmentation accuracy, a subset of smaller, anatomically accurate ROIs from the xtract ^37^ library was used, and binary masks were automatically generated for each ROI.

Diffusion MRI parameters were extracted within six white matter regions of interest: optic radiation (OR), anterior thalamic radiation (ATR), corticospinal tract (CST), forceps major (FMa), forceps minor (FMi), and brainstem (Bs). Bilateral tracts were analyzed as a single ROI by combining left and right masks. Tracts were chosen as they represent major projection, association, and commissural pathways, frequently implicated in MS-related disability ^11,38^. For each ROI, mean values were calculated. To assess changes in lesional WM and NAWM, ROI metrics were extracted before and after the exclusion of voxels corresponding to white matter lesions. Lesion masks were segmented by Samseg in FLAIR geometric space and later aligned to the diffusion space using *mri_easyreg*, a deep learning-based registration tool ^39^. This registration was performed using the spherical mean at b=2000 as the reference image in the diffusion space due to its contrast similarity with FLAIR, enabling accurate alignment. All segmentations were visually inspected by a neuroradiologist with 7 years of experience (VS).

### Statistical Methods

Spearman correlation analysis was conducted to assess the relationships between dMRI metrics and clinical measures across the selected ROIs. A p-value of <0.05 was considered statistically significant for all tests, with multiple comparisons corrected using the Bonferroni method.

Sex and age at disease onset were included as covariates to account for their established role in shaping disability progression in MS such as older age at onset being linked to faster accumulation of disability ^40^ as well as the female predominance of the disease ^41^. Since age and MS progression are closely connected, we aimed to account for the effect of aging, distinguishing it from the effects of the disease itself. Therefore, we adjusted for age at disease onset rather than overall age to better isolate disease-related changes.

All statistical analyses were performed using MATLAB v9.13.0 (R2022b) (The MathWorks Inc., Natick, MA).

## RESULTS

Figure 1 shows the fraction of voxels within the WM that correlate with each clinical metric (SDMT, 9HPT, MSFC, EDSS, and Disease Duration). SDMT exhibits the largest percentage of significant voxels across diffusion metrics, whereas EDSS shows the fewest. For all imaging metrics, most significant correlations are located in normal-appearing white matter (NAWM), with only a smaller fraction overlapping with lesions. In the following sections, we describe for each clinical outcome the voxelwise and ROI-based correlations with diffusion metrics and compare them to conventional volumetric measures and lesion load. We focus on RD and RK, as these metrics have been shown to be the most sensitive to demyelination ^18,42^, and on the SMI metrics *f* and *p_2_*, selected as the most reliable parameters obtainable from two-shell protocols, as outlined in the Methods.

**Figure 1.**
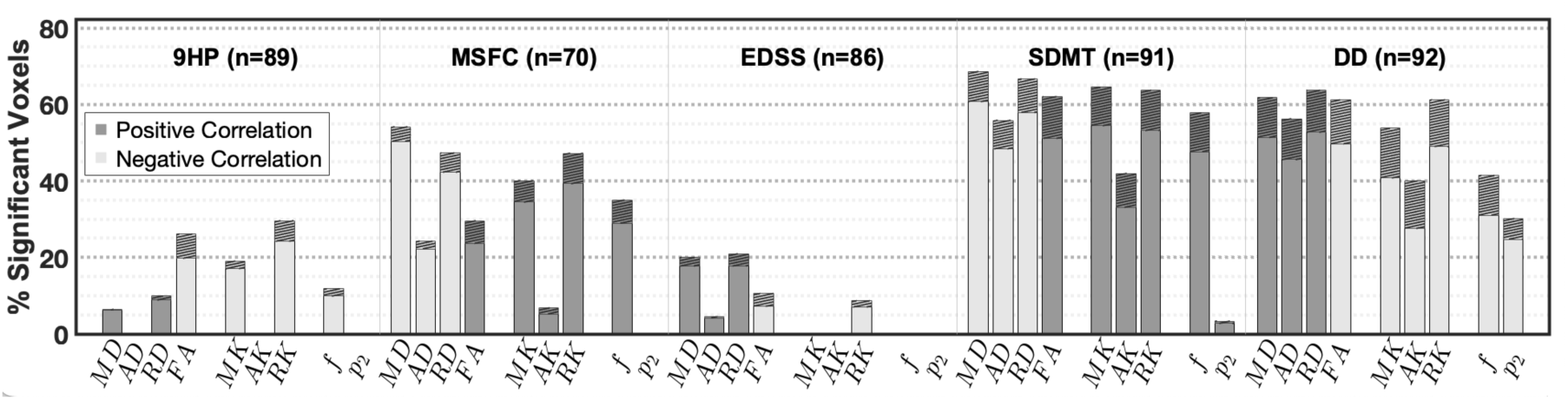
Sensitivity of diffusion metrics to MS disability. For each clinical measure (9-Hole Peg Test (9HPT); Multiple Sclerosis Functional Composite (MSFC); Expanded Disability Status Scale (EDSS); Symbol Digit Modalities Test (SDMT); and Disease Duration (DD) and each group of diffusion metrics (Diffusion Tensor Imaging (DTI); Diffusion Kurtosis Imaging (DKI); Standard Model Imaging, (SMI), the bars show the percentage of white matter voxels with significant correlations (p < 0.05) between diffusion metrics and clinical scores. Dark gray bars represent positive correlations, light gray bars represent negative correlations. The hatched upper segment of each bar indicates the fraction of significant voxels overlapping with the lesion probability map (voxels lesional in ≥10% of subjects).

### Correlation with 9HPT

Voxelwise analysis revealed tract-specific correlations with 9HPT performance, in agreement with ROI-based results (Figure 2). The strongest correlations were found in the CST (RD: ρ = 0.45, RK: ρ = −0.44, *f*: ρ = −0.39), consistent with its role in motor control. The ATR also correlated (RD: ρ = 0.44, RK: ρ = −0.37, *f*: ρ = −0.39), supporting thalamo-frontal contributions to attentional control, while the OR showed correlations (RD: ρ = 0.40, RK: ρ = −0.43, *f*: ρ = −0.42), reflecting visuomotor integration during the task. The brainstem correlated (RD: ρ = 0.37, RK: ρ = −0.42, *f*: ρ = −0.40), in line with its role as the conduit for corticospinal and cerebellar pathways.

**Figure 2.**
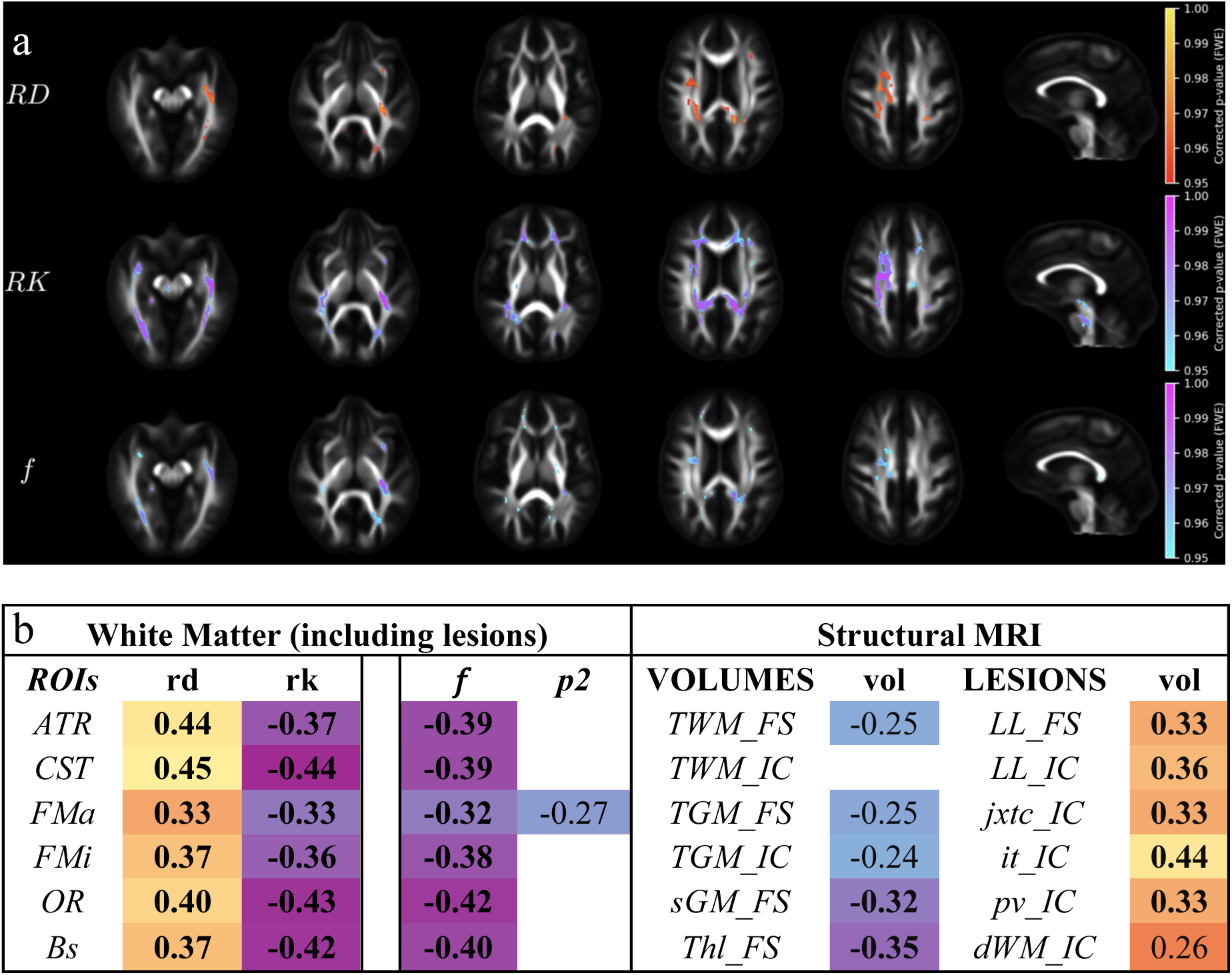
Correlations between imaging metrics and 9-Hole Peg Test (9HPT). **(a)** Voxel-wise correlations between diffusion metrics and 9HPT. Significant clusters (p ≤ 0.05, FWE-corrected) are shown within a white matter mask (FA > 0.2). Warm colors indicate positive correlations; cool colors indicate negative correlations. Results are displayed for radial diffusivity (RD), radial kurtosis (RK), and intra-axonal volume fraction (*f*). **(b)** ROI-based correlations between 9HPT and imaging measures (dMRI metrics in WM and structural volumes). Spearman’s ρ values are shown; those surviving Bonferroni correction are in bold. Abbreviations: ATR, anterior thalamic radiation; CST, corticospinal tract; FMa, forceps major; FMi, forceps minor; OR, optic radiation; BS, brainstem; TWM, total WM; TGM, total GM; sGM, subcortical GM; Thl, thalamus; LL, lesion load; FS, FreeSurfer; IC, Icometrix; jc, juxtacortical; inf, infratentorial; pv, periventricular; dWM, deep WM.

Excluding lesions had little impact, with ROI-based correlations remaining highly similar (see Supplementary Figure 1). The largest changes were modest, such as in the CST (RD: ρ = 0.45 → 0.47) and OR (RD: ρ = 0.40 → 0.42), while correlations in other tracts varied minimally. This confirms that correlations are largely driven by NAWM.

By comparison, structural MRI showed weaker effects. Among brain volumes, the most consistent correlate was thalamus volume (ρ = −0.35). Among lesion metrics, infratentorial lesion load showed the strongest structural association (ρ = 0.44), indicating the importance of cerebellar and brainstem lesions in motor impairment.

All correlations reported here remained significant after Bonferroni correction.

### Correlation with MSFC

Voxelwise analysis revealed widespread correlations with MSFC performance, in agreement with ROI-based results (Figure 3). The strongest associations were observed in the brainstem (RD: ρ = −0.52, RK: ρ = 0.44, *f*: ρ = 0.42), consistent with its integrative role in corticospinal and cerebellar pathways supporting gait and hand function. The optic radiations also showed strong correlations (RD: ρ = −0.41, RK: ρ = 0.40, *f*: ρ = 0.39), reflecting the visual contributions across tasks. Additional associations were seen in the ATR (RD: ρ = −0.37) and CST (RD: ρ = −0.34) showed weaker correlations, supporting thalamo-frontal attentional control and motor conduction, respectively.

**Figure 3.**
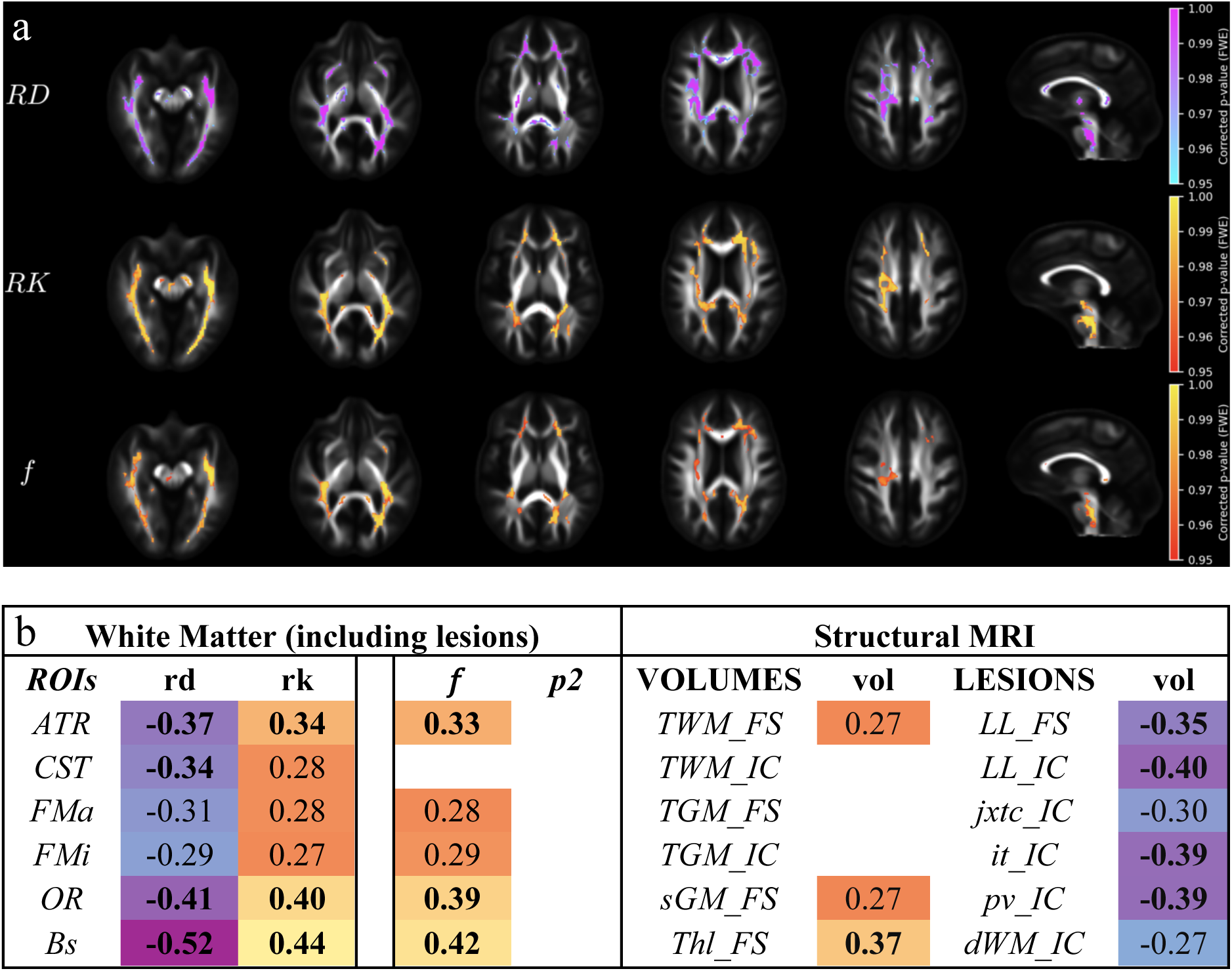
Correlations between imaging metrics and Multiple Sclerosis Functional Composite (MSFC). **(a)** Voxel-wise correlations between diffusion metrics and MSFC. Significant clusters (p ≤ 0.05, FWE-corrected) are shown within a white matter mask (FA > 0.2). Warm colors indicate positive correlations; cool colors indicate negative correlations. Results are displayed for radial diffusivity (RD), radial kurtosis (RK), and intra-axonal volume fraction (*f*). **(b)** ROI-based correlations between MSFC and imaging measures (dMRI metrics in WM and structural volumes). Spearman’s ρ values are shown; those surviving Bonferroni correction are in bold. Abbreviations: ATR, anterior thalamic radiation; CST, corticospinal tract; FMa, forceps major; FMi, forceps minor; OR, optic radiation; BS, brainstem; TWM, total white matter; TGM, total gray matter; sGM, subcortical gray matter; Thl, thalamus; LL, lesion load; jxtc, juxtacortical; it, infratentorial; pv, periventricular; dWM, deep white matter; FS, FreeSurfer; IC, Icobrain.

Excluding lesions had little effect, with ROI-based correlations remaining highly similar (see Supplementary Figure 2). For example, the OR maintained strong associations (*f*: ρ = 0.39 → 0.38). Overall, these findings indicate that MSFC associations are largely driven by NAWM.

Structural measures showed weaker associations. Among brain volumes, thalamus (ρ = 0.37) correlated modestly. Among lesion metrics, total lesion load had the highest association (ρ = −0.40), followed by periventricular and infratentorial and lesion loads (each ρ = −0.39), underscoring the contribution of cerebellar and brainstem lesions to MSFC impairment.

All correlations reported here remained significant after Bonferroni correction.

### Correlation with EDSS

Voxelwise analysis revealed fewer and more scattered correlations compared to 9HPT and MSFC; however, ROI-based analysis (Figure 4) showed correlations spanning most ROIs, though weaker than those observed for 9HPT and MSFC. The strongest diffusion correlations were found in the ATR (RD: ρ = 0.37, *f*: ρ = −0.32) and CST (RD: ρ = 0.32, RK: ρ = −0.32), consistent with their established roles in motor disability. Importantly, correlations also appeared in commissural fibers (FMa, FMi) with ***p_2_*** (ρ = −0.32, −0.30), indicating reduced fiber alignment likely linked to chronic inflammatory changes. This marks the first appearance of commissural disorganization in a motor/disability measure, providing a bridge to patterns observed with Disease Duration. Notably, no significant diffusion correlations were detected in the brainstem, marking a shift from the tract-specific associations seen in functional measures.

**Figure 4.**
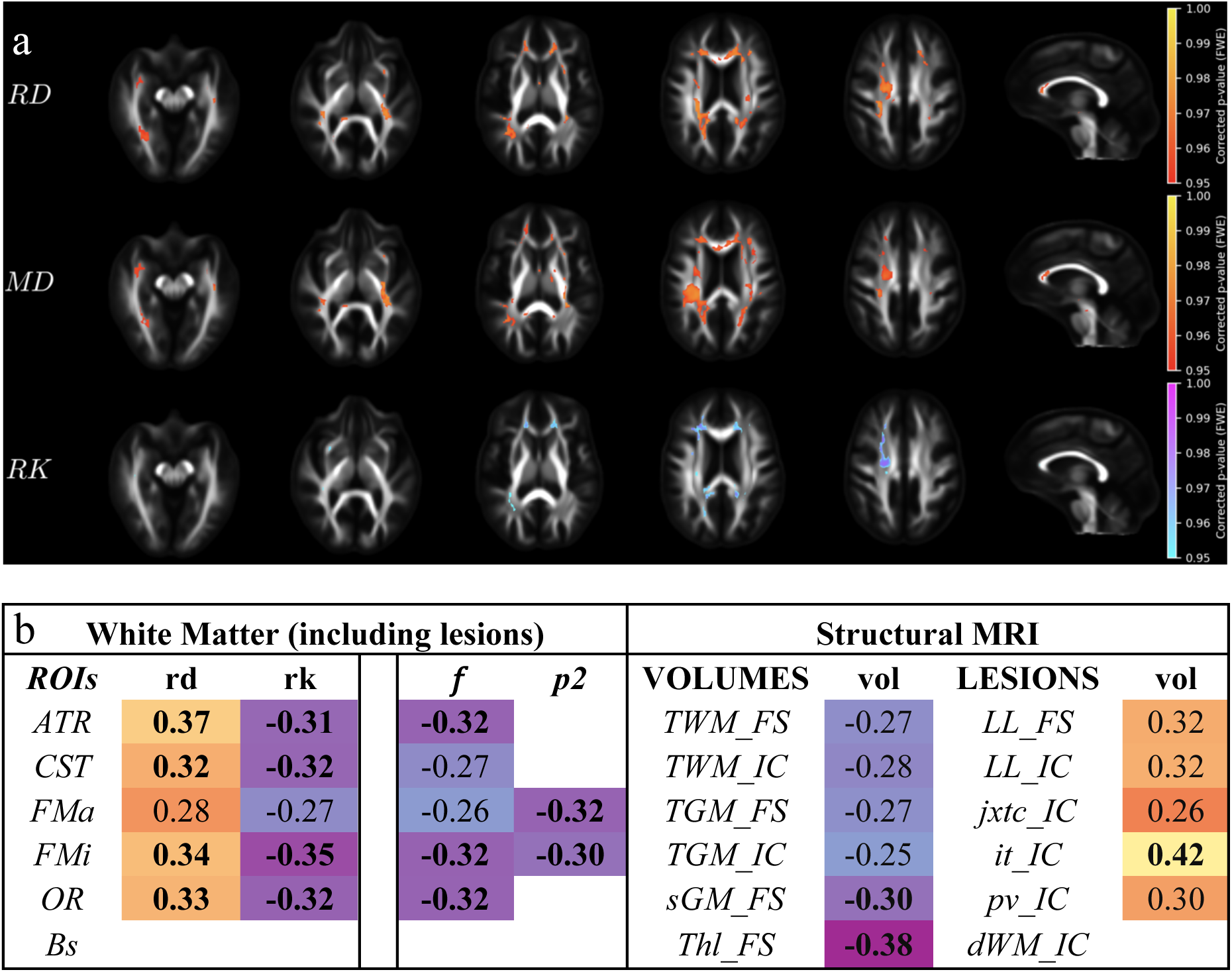
Correlations between imaging metrics and Expanded Disability Status Scale (EDSS). **(a)** Voxel-wise correlations between diffusion metrics and EDSS. Significant clusters (p ≤ 0.05, FWE-corrected) are shown within a white matter mask (FA > 0.2). Warm colors indicate positive correlations; cool colors indicate negative correlations. Results are displayed for radial diffusivity (RD), mean diffusivity (MD), and radial kurtosis (RK). **(b)** ROI-based correlations between EDSS and imaging measures (dMRI metrics in WM and structural volumes). Spearman’s ρ values are shown; those surviving Bonferroni correction are in bold. Abbreviations: ATR, anterior thalamic radiation; CST, corticospinal tract; FMa, forceps major; FMi, forceps minor; OR, optic radiation; BS, brainstem; TWM, total white matter; TGM, total gray matter; sGM, subcortical gray matter; Thl, thalamus; LL, lesion load; jxtc, juxtacortical; it, infratentorial; pv, periventricular; dWM, deep white matter; FS, FreeSurfer; IC, Icobrain.

Excluding lesions had little effect, with ROI-based correlations remaining largely stable (see Supplementary Figure 3). For example, CST (RD: ρ = 0.32 → 0.31; RK: ρ = −0.32 → −0.35), ATR (RD: ρ = 0.37 → 0.37; RK: ρ = −0.31 → −0.33). Overall, these findings confirm that EDSS associations are largely driven by NAWM. All correlations reported here remained significant after Bonferroni correction.

Among structural measures, thalamus atrophy emerged as the dominant volumetric correlate (ρ = −0.38), highlighting its role as a marker of late-stage degeneration. In addition, infratentorial lesion load showed the strongest overall correlation (ρ = 0.42), underscoring the contribution of cerebellar and brainstem lesions to motor disability, as consistently observed across all motor-related measures.

Together, these features distinguish EDSS from functional performance measures such as 9HPT and MSFC and align it more closely with the cumulative pattern observed for Disease Duration.

### Correlation with Cognitive Performance (SDMT)

Voxelwise analysis revealed widespread correlations with SDMT, which were mirrored in ROI-based results (Figure 5). Diffusion metrics showed strong associations across nearly all ROIs, for example in the ATR (RD: ρ = −0.43, RK: ρ = 0.39, *f*: ρ = 0.40), CST (RD: ρ = −0.40, RK: ρ = 0.38, *f*: ρ = 0.39), and OR (RD: ρ = −0.42, RK: ρ = 0.43, *f*: ρ = 0.43). The intra-axonal water fraction (*f*) was most strongly associated with the OR, consistent with visual pathway involvement highlighted across multiple functional tests.

**Figure 5.**
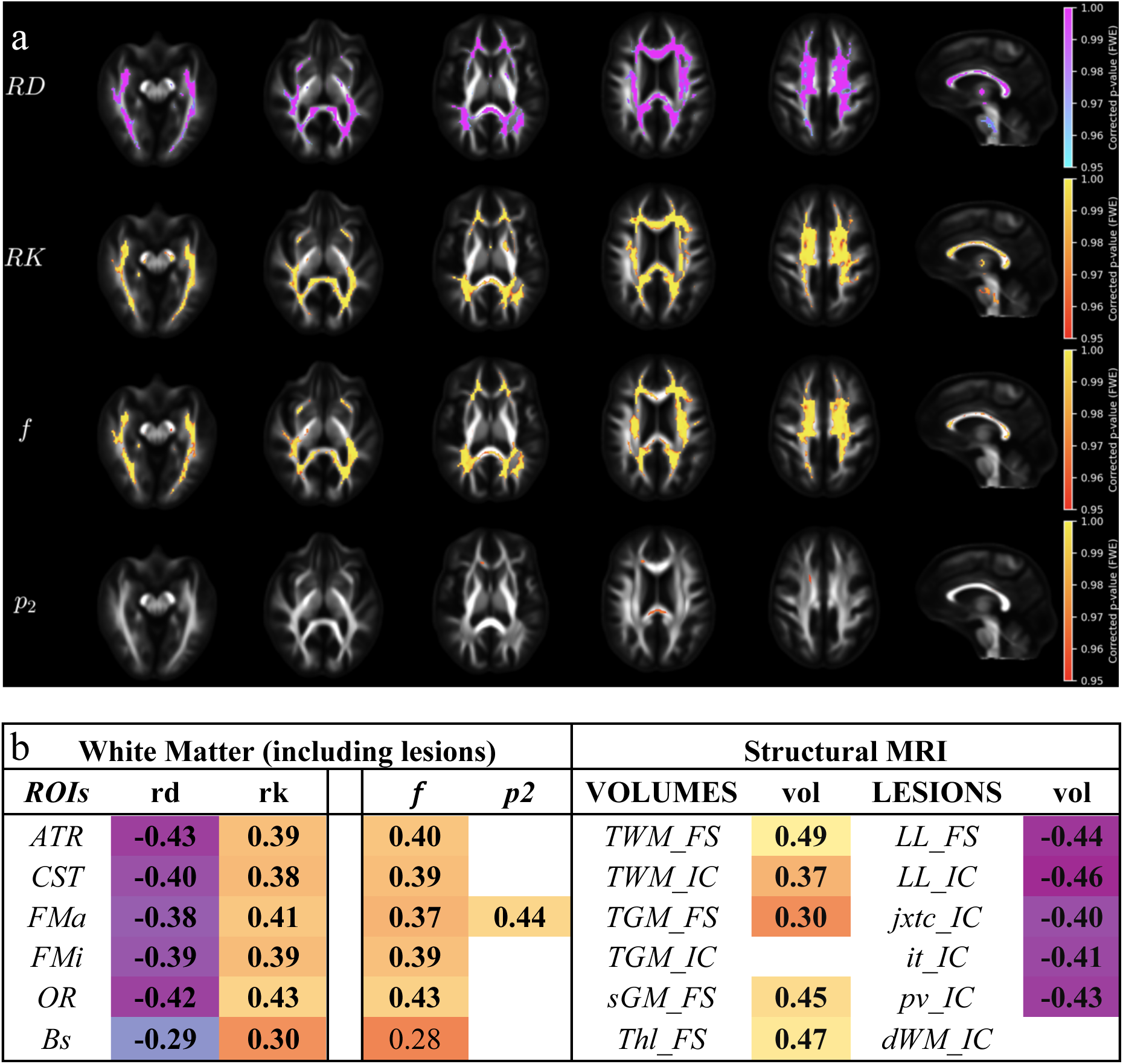
Correlations between imaging metrics and Symbol Digit Modalities Test (SDMT). **(a)** Voxel-wise correlations between diffusion metrics and SDMT. Significant clusters (p ≤ 0.05, FWE-corrected) are shown within a white matter mask (FA > 0.2). Warm colors indicate positive correlations; cool colors indicate negative correlations. Results are displayed for radial diffusivity (RD), radial kurtosis (RK), intra-axonal volume fraction (*f*), and fiber dispersion index (*p_2_*). **(b)** ROI-based correlations between SDMT and imaging measures (dMRI metrics in WM and structural volumes). Spearman’s ρ values are shown; those surviving Bonferroni correction are in bold. Abbreviations: ATR, anterior thalamic radiation; CST, corticospinal tract; FMa, forceps major; FMi, forceps minor; OR, optic radiation; BS, brainstem; TWM, total white matter; TGM, total gray matter; sGM, subcortical gray matter; Thl, thalamus; LL, lesion load; jxtc, juxtacortical; it, infratentorial; pv, periventricular; dWM, deep white matter; FS, FreeSurfer; IC, Icobrain.

This global pattern might be caused by the multifactorial nature of SDMT, which relies on visual, attentional, memory, and motor components rather than a single functional system.

Excluding lesions had a clear impact on the associations with SDMT (see Supplementary Figure 4). The largest change was observed in the ATR, where RD decreased from ρ = −0.43 to ρ = −0.37. Additional decreases were noted in FMa (RK: ρ = 0.41 to 0.38; *f*: ρ = 0.37 to 0.33) and OR (*f*: ρ = 0.43 to 0.39). These results indicate that, compared to other clinical measures, SDMT correlations are more affected by lesion exclusion, though NAWM pathology still accounts for the majority of associations.

Structural MRI measures similarly demonstrated strong correlations, with total WM (ρ = 0.49), thalamus (ρ = 0.47), and subcortical GM (ρ = 0.45) among the top volumetric measures. Lesion metrics also showed consistent correlations across categories (ρ = −0.40 to −0.46).

Together, these results show that SDMT performance is strongly associated with both WM microstructural integrity and GM/WM atrophy, but without a single dominant spatial pattern.

### Correlation with Disease Duration

Voxelwise analysis revealed widespread correlations, whereas ROI-based analysis (Figure 6) showed more modest associations. The most consistent finding was in the commissural fibers, with reduced *p_2_* in the FMa (ρ = −0.45) and FMi (ρ = −0.43), which indicates fiber misalignment that may be related to long-term disease effects. This commissural involvement mirrors the EDSS pattern and highlights a shared signature of cumulative burden. Additional correlations were observed in the ATR (RD: ρ = 0.37) and OR (RD: ρ = 0.33), though these were weaker than those linked to functional measures. No significant correlations were detected in the brainstem.

**Figure 6.**
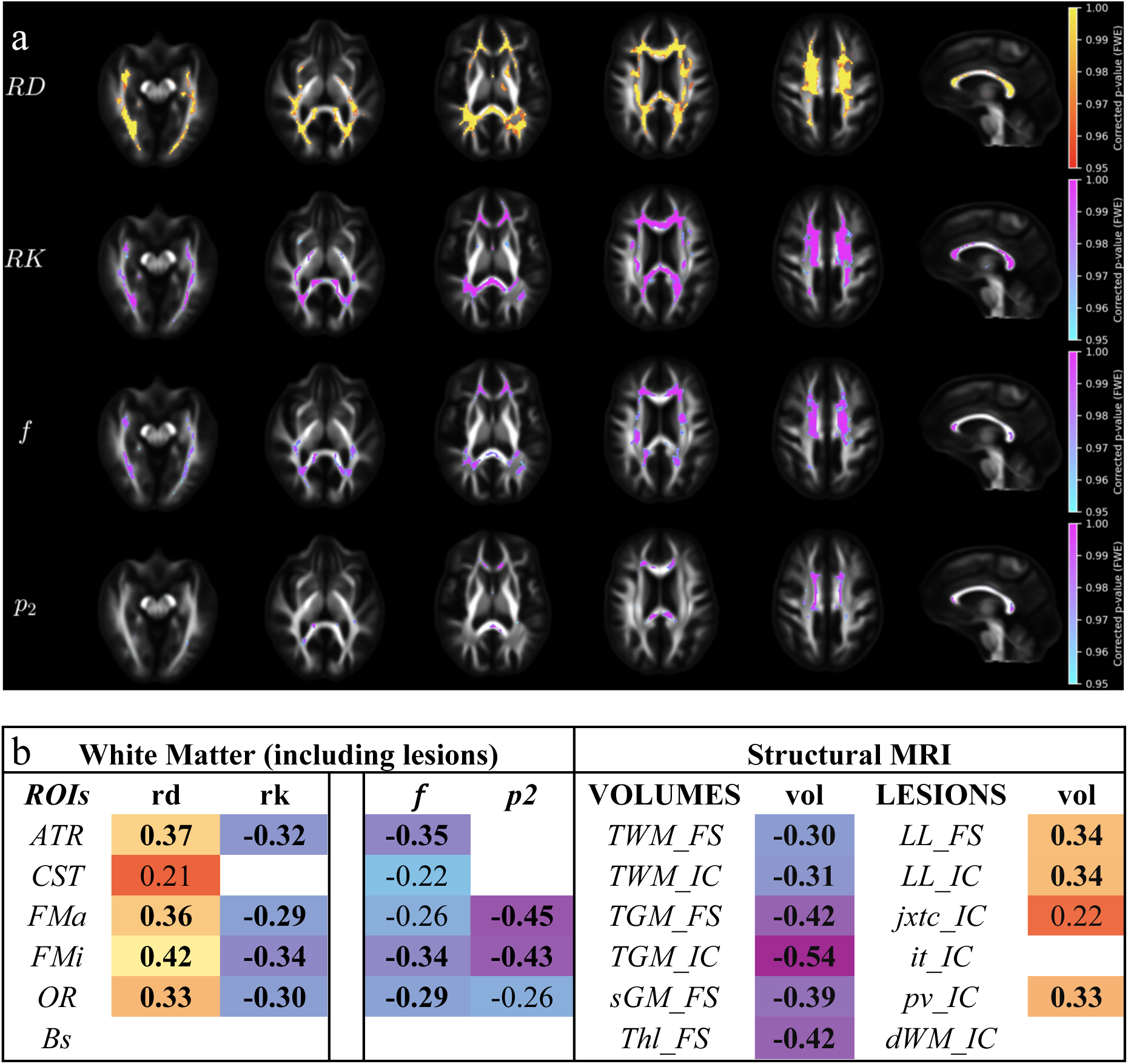
Correlations between imaging metrics and disease duration. **(a)** Voxel-wise correlations between diffusion metrics and disease duration. Significant clusters (p ≤ 0.05, FWE-corrected) are shown within a white matter mask (FA > 0.2). Warm colors indicate positive correlations; cool colors indicate negative correlations. Results are displayed for radial diffusivity (RD), radial kurtosis (RK), intra-axonal volume fraction (*f*), and fiber dispersion index (*p_2_*). **(b)** ROI-based correlations between disease duration and imaging measures (dMRI metrics in WM and structural volumes). Spearman’s ρ values are shown; those surviving Bonferroni correction are in bold. Abbreviations: ATR, anterior thalamic radiation; CST, corticospinal tract; FMa, forceps major; FMi, forceps minor; OR, optic radiation; BS, brainstem; TWM, total white matter; TGM, total gray matter; sGM, subcortical gray matter; Thl, thalamus; LL, lesion load; jxtc, juxtacortical; it, infratentorial; pv, periventricular; dWM, deep white matter; FS, FreeSurfer; IC, Icobrain.

Excluding lesions had little impact and slightly reduced the strength of commissural effects (see Supplementary Figure 5): *p_2_* in FMa (ρ = −0.45 vs −0.40) and FMi (ρ = −0.43 vs −0.40). Smaller changes were seen for ATR (RD: ρ = 0.37 vs 0.34) and OR (RD: ρ = 0.33 vs 0.32). These results confirm that disease-duration associations are largely driven by NAWM.

Structural measures showed stronger effects, dominated by gray matter atrophy, most notably total GM (ρ = −0.54), followed by the thalamus (ρ = −0.42) and subcortical GM (ρ = −0.39). WM volumes showed weaker associations, and lesion metrics were modest, with no contribution from infratentorial lesions. All correlations reported here remained significant after Bonferroni correction.

## DISCUSSION

This study systematically compared advanced diffusion MRI (DTI, DKI, SMI), volumetrics, and lesion metrics in MS. dMRI metrics are calculated within each voxel and are by design not directly sensitive to pseudoatrophy after therapy^5,43^. We integrated voxelwise and ROI-based approaches across NAWM and lesional tissue. Unlike prior studies ^10,11,13^ that focused on a single modality, we combined multiple quantitative methods to link microstructural, structural, and regional lesion metrics to clinical outcomes. Novel aspects include the incorporation of SMI parameters, which move beyond empirical diffusion signal representations (DTI, DKI) to biologically interpretable markers, and the quantification of both global and regional lesion load ^25,26^, which proved more clinically informative than global lesion metrics.

Figure 1 provides an overview of the voxelwise associations between diffusion metrics and clinical outcomes, highlighting both their extent and distribution across WM. SDMT exhibited the largest percentage of significant voxels, whereas EDSS showed the fewest. Importantly, most correlations were located in NAWM, with only a smaller fraction overlapping lesions. ROI-based analyses (Figures 2-6 (b) and Supplementary Figures 1-5) further showed that correlations were preserved after excluding lesional voxels, with only minimal changes in effect sizes, indicating that lesion burden alone does not account for clinical variability. This underscores NAWM pathology as a key contributor to functional impairment, even in tissue that appears normal on conventional MRI. These findings are consistent with prior work demonstrating diffusion abnormalities in NAWM^13,44,45^.

DTI- and DKI-derived measures showed robust, tract-specific associations with motor and cognitive outcomes, largely driven by NAWM pathology. While for exploratory voxelwise analysis we initially considered all DTI/DKI (Figure 1), we then focused on RD and RK for ROI-based analysis (Figures 2-6, Tables 2-6) as these are the most sensitive to demyelination ^18,42,46^. Functional measures (9HPT, MSFC, SDMT) exhibited the strongest and most spatially specific diffusion associations, with tract involvement in CST, ATR, OR, and brainstem. By contrast, EDSS and disease duration displayed weaker and less localized diffusion correlations, which are considered in more detail below.

Both DTI (e.g., RD) and DKI (e.g., RK) metrics showed consistent correlations with clinical outcomes. In several regions, particularly the optic radiations for SDMT, RK provided slightly stronger associations, while in other tracts RD was equally or more robust. Overall, the two approaches demonstrated highly convergent patterns, suggesting that DKI offers complementary sensitivity rather than a uniform advantage. This complementarity is consistent with prior work showing that DKI detects NAWM abnormalities and microstructural complexity not fully captured by tensor metrics ^47^. An additional advantage of multi-shell acquisitions is that they also enable extraction of biophysically grounded parameters such as those derived from SMI.

SMI parameters added mechanistic specificity, with the intra-axonal water fraction (*f*) probing axonal loss and demyelination in the CST, ATR, and OR, linking motor and visuomotor pathway damage to 9HPT and MSFC performance. For cognitive function (SDMT), *f* was most strongly associated with the optic radiations, consistent with the visual dependence of this task. For disability (EDSS), *f* correlated with the ATR, FMi, and OR, indicating that axonal loss in these tracts contributes to motor impairment. The fiber dispersion index (*p_2_*) identified commissural disorganization in the FMa and FMi, associated with both EDSS and disease duration, consistent with the voxelwise patterns in Figures 4 and 6, and suggesting inflammation-related misalignment of highly ordered callosal fibers. These parameters have been validated against histology in animal models ^18,19,42^, supporting their interpretation as biologically grounded markers of microstructural pathology.

In interpreting the MRI associations, we noted that the outcome measures could be grouped based on both their clinical nature and the patterns of correlations observed. EDSS and disease duration showed broad, non-specific diffusion associations, consistent with cumulative burden. In contrast, MSFC and 9HPT, composed of timed performance tasks, displayed more focal, tract-specific associations, justifying their classification as ‘functional’ outcomes. SDMT showed distributed associations across diffusion and structural metrics, in line with its multifactorial demands on processing speed, attention, and motor output. These distinctions motivated the use of terms such as “cumulative disease burden” for EDSS and disease duration, and “functional” for MSFC and 9HPT. ROI analyses further distinguished cumulative disease burden outcomes: EDSS and disease duration were both linked to commissural disorganization (*p_2_* reductions in FMa/FMi) and gray matter atrophy but differed in emphasis. EDSS retained motor-specific correlations (CST, ATR, OR) and was strongly tied to infratentorial lesion load, underscoring brainstem and cerebellar involvement. Disease duration instead showed stronger associations with global and thalamic gray matter atrophy, with weaker tract or infratentorial effects. Thus, EDSS reflects both motor disability, whereas disease duration primarily tracks long-term structural damage. Structural measures were less strongly associated with motor performance but captured cumulative effects. Thalamic atrophy was the most consistent volumetric correlate, particularly for SDMT and disease duration, in line with its established role as a marker of MS progression ^48^. Subcortical GM and total WM volumes also contributed, especially for SDMT and duration, consistent with widespread tissue loss driving cognitive and global impairment.

Regional lesion load provided stronger clinical associations than global lesion burden. For 9HPT, infratentorial lesion load was the strongest structural correlate, aligning with the clinical role of brainstem and cerebellar lesions in motor coordination. For MSFC, total lesion load correlated, with periventricular and infratentorial lesions. For EDSS, infratentorial lesion load again emerged as the strongest structural correlate, underscoring its role in motor disability. In contrast, disease duration correlated with cortical and subcortical GM atrophy, but not with lesion load. These results are consistent with earlier work showing that infratentorial and periventricular lesions disproportionately contribute to disability ^49,50^, underscoring the value of regional over global lesion mapping.

Direct comparison of modalities (diffusion vs structural MRI) revealed a consistent pattern across outcomes. DTI and DKI metrics were most sensitive to functional measures such as 9HPT and MSFC, where tract-specific associations in CST, ATR, OR, and brainstem predominated. In contrast, structural measures - particularly thalamic and global GM atrophy - showed stronger correlations with disease duration. For SDMT, the strongest structural associations (ρ up to 0.49 for total WM, 0.47 for thalamus) modestly exceeded those observed with DTI/DKI (ρ up to 0.43 in OR and ATR), suggesting that cognitive performance is more strongly linked to volumetric atrophy, while diffusion highlights distributed WM contributions. EDSS displayed an intermediate profile: DTI/DKI highlighted commissural fiber disorganization, whereas infratentorial lesion load and thalamic atrophy emerged as the strongest structural correlates. These tract-specific associations were observed consistently across both voxelwise maps and ROI-based analyses (**Figures 2-6**).

These findings support the hypothesis that microstructural degeneration in NAWM precedes measurable atrophy, with diffusion abnormalities explaining functional impairment, while volumetric atrophy and lesion burden reflect cumulative damage. This temporal sequence provides a unifying framework for interpreting the complementary roles of diffusion and structural imaging in MS. These results suggest that microstructural changes may precede measurable atrophy, although longitudinal studies are needed to confirm this sequence.

This study has several limitations. First, its cross-sectional design precludes definitive conclusions about the temporal sequence of diffusion abnormalities and atrophy, which require longitudinal validation. Second, while SMI parameters provide biologically interpretable markers of axonal loss and fiber disorganization, they remain model-based estimates without direct histological validation in patients; however, these measures have been validated in animal models, including the cuprizone mouse model ^18,42^. Third, the study applied a protocol optimized for routine clinical use, balancing acquisition time and parameter estimation. While this approach enables estimation of the Standard Model parameters *f* and *p*_2_, it does not support inclusion of free water as a separate compartment, which would be possible with expanded acquisition protocols in future work^8^. A further limitation concerns the estimation of lesion burden, which depends on the processing pipeline, though was evaluated by using two commonly used software tools ^26^. While results were consistent for global lesion measures across two segmentation approaches, only one method allowed regional quantification, which improved clinical associations. Finally, this study did not include spinal cord imaging, which is tightly related to disability and could have added further clinical relevance.

**In conclusion,** this study shows that diffusion MRI captures clinically meaningful microstructural abnormalities in MS, extending beyond focal lesions and highlighting NAWM damage as a key driver of disability. Diffusion metrics demonstrated stronger and more spatially specific associations with functional outcomes, while structural measures such as thalamic atrophy and infratentorial lesion load related more to accumulated disease burden captured by EDSS and disease duration. Standard Model Imaging adds biological specificity, with the intra-axonal fraction (*f*) reflecting axonal loss and demyelination and the fiber dispersion index (*p_2_*) capturing commissural disorganization linked to cumulative burden. Taken together, these findings highlight the complementary value of combining diffusion with volumetric and lesion-based measures. Clinically feasible multi-shell diffusion protocols may therefore improve monitoring and provide a more comprehensive framework for understanding and managing disease progression in MS.

## Data Availability

All data produced in the present study are available upon reasonable request to the authors

## List of Abbreviations

MS: Multiple Sclerosis
WM: White Matter
GM: Gray Matter
dMRI: diffusion MRI
LL: Lesion Load
DTI: Diffusion Tensor Imaging
DKI: Diffusion Kurtosis Imaging
SMI: Standard Model Imaging
SDMT: Symbol Digit Modalities Test
EDSS: Expanded Disability Status Scale
MSFC: Multiple Sclerosis Functional Composite
ROI: Region of Interest
CST: Corticospinal Tract
OR: Optic Radiation
NAWM: Normal-Appearing White Matter
*f*: Intra-axonal Volume Fraction
*p2*: Orientation Dispersion Index
SM: Standard Model
ODF: Orientation Distribution Function
MPRAGE: Magnetization Prepared Rapid Gradient Echo
TR: Repetition Time
TI: Inversion Time
TE: Echo Time
FA: Fractional Anisotropy
TA: Total Acquisition Time
FLAIR: Fluid-Attenuated Inversion Recovery
EPI: Echo Planar Imaging
GRAPPA: Generalized Autocalibrating Partial Parallel Acquisition
TIV: Total Intracranial Volume
VS: Voxel Size
WLLS: Weighted Linear Least Squares
MD: Mean Diffusivity
AD: Axial Diffusivity
RD: Radial Diffusivity
MK: Mean Kurtosis
AK: Axial Kurtosis
RK: Radial Kurtosis
TFCE: Threshold-Free Cluster Enhancement
FWE: Family-Wise Error
DD: Disease Duration
ATR: Anterior Thalamic Radiation
FMa: Forceps Major
FMi: Forceps Minor
Bs: Brainstem
9HPT: 9-Hole Peg Test

## Credit authorship contribution statement

Valentin N. Stepanov: Writing - original draft, Visualization, Conceptualization, Formal analysis. Santiago Coelho: Software, Writing - review & editing, Conceptualization. Benjamin Ades-Aron: Software. Jenny Chen: Software. Timothy M. Shepherd: Writing - review & editing, Supervision, Conceptualization. Ilya Kister: Patient recruitment, Writing - review & editing, Supervision, Conceptualization. Dmitry S. Novikov: Writing - review & editing, Supervision, Conceptualization. Els Fieremans: Writing - review & editing, Supervision, Conceptualization.

## Acknowledgments

We thank Nalini Jeet, Carli Salvati, Fade Djemo, Nadine Azmy, and Francesca Chinea for participant recruitment. We also acknowledge Mary Bruno and Wafaa Sweidan for assistance with data acquisition.

## Data and code availability

The datasets used and/or analyzed during the current study will be available from the corresponding author on reasonable request.

## Supplementary Information

**Table 1:**
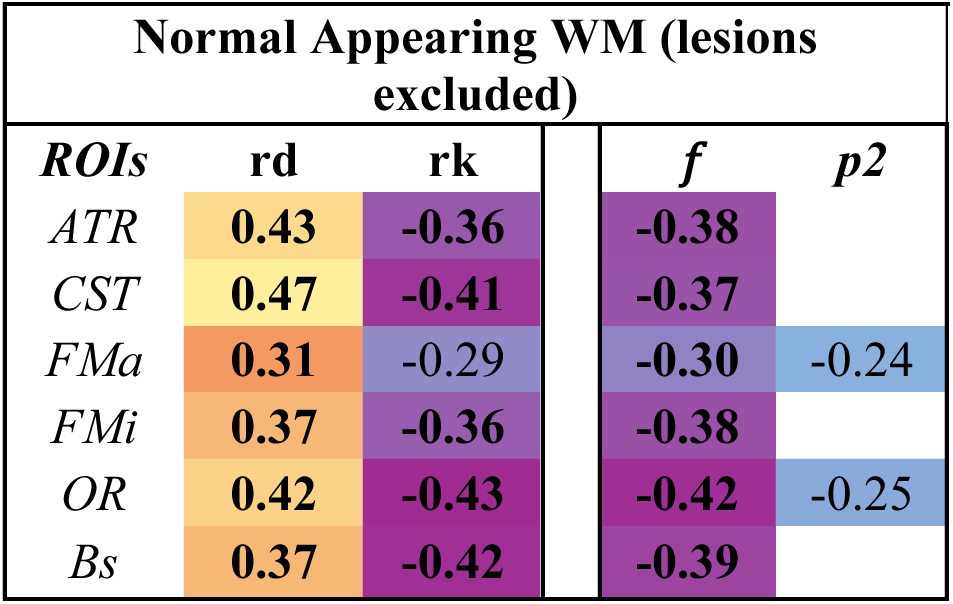
9-Hole Peg Test. W-Kurtosis, DTI-derived DTI maps Correlations between 9HPT and imaging measures after lesion exclusion (dMRI in NAWM and Structural Volumes). Note: ρ represents the Spearman correlation coefficient. Values that remained significant after Bonferroni correction are shown in bold. ATR, anterior thalamic radiation; CST, corticospinal tract; FMa, forceps major; FMi, forceps minor; OR, optic radiation; Bs, brainstem.

**Table 2:** MSFC. W-Kurtosis, DTI-derived DTI maps Correlations between MSFC and imaging measures after lesion exclusion (dMRI in NAWM and Structural Volumes). Note: ρ represents the Spearman correlation coefficient. Values that remained significant after Bonferroni correction are shown in bold. ATR, anterior thalamic radiation; CST, corticospinal tract; FMa, forceps major; FMi, forceps minor; OR, optic radiation; Bs, brainstem.

| Normal Appearing WM (lesions excluded) |  |  |  |  |
| --- | --- | --- | --- | --- |
| *ROIs* | *rd* | *rk* | *f* | *p<sup>2</sup>* |
| *ATR* | **-0.34** | 0.33 | 0.31 |  |
| *CST* | **-0.36** | 0.28 |  |  |
| *FMa* | -0.27 | 0.24 |  |  |
| *FMi* | -0.28 | 0.27 | 0.29 |  |
| *OR* | **-0.40** | **0.40** | **0.38** | 0.26 |
| *Bs* | **-0.52** | **0.44** | **0.42** |  |

**Table 3:** EDSS. W-Kurtosis, DTI-derived DTI maps Correlations between EDSS and imaging measures after lesion exclusion (dMRI in NAWM and Structural Volumes). Note: ρ represents the Spearman correlation coefficient. Values that remained significant after Bonferroni correction are shown in bold. ATR, anterior thalamic radiation; CST, corticospinal tract; FMa, forceps major; FMi, forceps minor; OR, optic radiation; Bs, brainstem.

| Normal Appearing WM (lesions excluded) |  |  |  |  |  |
| --- | --- | --- | --- | --- | --- |
| *ROIs* | *rd* | *rk* |  | *f* | *p2* |
| *ATR* | **0.37** | **-0.33** |  | -0.29 |  |
| *CST* | **0.31** | **-0.35** |  | -0.22 |  |
| *FMa* | 0.25 | -0.24 |  | -0.21 | **-0.31** |
| *FMi* | **0.32** | **-0.36** |  | **-0.33** | -0.28 |
| *OR* | **0.31** | **-0.31** |  | -0.29 |  |
| *Bs* |  |  |  |  |  |

**Table 4:** SDMT. W-Kurtosis, DTI-derived DTI maps Correlations between SDMT and imaging measures after lesion exclusion (dMRI in NAWM and Structural Volumes). Note: ρ represents the Spearman correlation coefficient. Values that remained significant after Bonferroni correction are shown in bold. ATR, anterior thalamic radiation; CST, corticospinal tract; FMa, forceps major; FMi, forceps minor; OR, optic radiation; Bs, brainstem.

| Normal Appearing WM (lesions excluded) |  |  |  |  |  |
| --- | --- | --- | --- | --- | --- |
| *ROIs* | *rd* | *rk* |  | *f* | *p2* |
| *ATR* | **-0.37** | **0.35** |  | **0.36** | **-0.23** |
| *CST* | **-0.39** | **0.35** |  | **0.35** |  |
| *FMa* | **-0.36** | **0.38** |  | **0.33** | **0.43** |
| *FMi* | **-0.36** | **0.37** |  | **0.38** |  |
| *OR* | **-0.44** | **0.42** |  | **0.39** | **0.32** |
| *Bs* | **-0.29** | **0.30** |  | 0.28 |  |

**Table 5:**
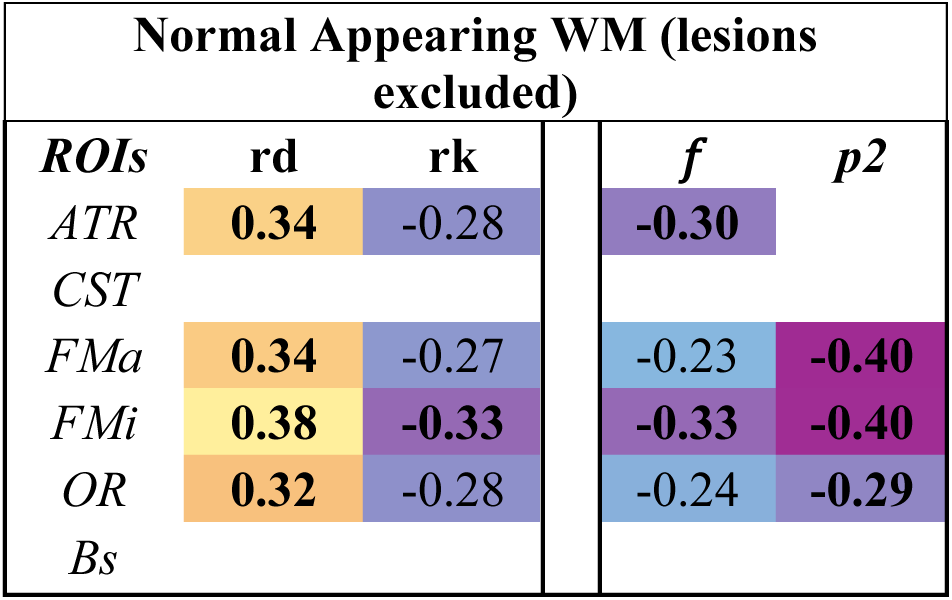
Disease Duration. W-Kurtosis, DTI-derived DTI maps Correlations between disease duration and imaging measures after lesion exclusion (dMRI in NAWM and Structural Volumes). Note: ρ represents the Spearman correlation coefficient. Values that remained significant after Bonferroni correction are shown in bold. ATR, anterior thalamic radiation; CST, corticospinal tract; FMa, forceps major; FMi, forceps minor; OR, optic radiation; Bs, brainstem.

